# Resistance to cortical amyloid-beta associates with cognitive health in centenarians

**DOI:** 10.1101/2023.12.28.23300604

**Authors:** Susan K. Rohde, Patricia Fierro-Hernández, Annemieke J.M. Rozemuller, Netherlands Brain Bank, Linda M.C. Lorenz, Meng Zhang, Marieke Graat, Myke van der Hoorn, Dominique Daatselaar, Marc Hulsman, Philip Scheltens, Sietske A.M. Sikkes, Jeroen J.M. Hoozemans, Henne Holstege

**Affiliations:** Department of Human Genetics, Genomics of Neurodegenerative Diseases and Aging, Vrije Universiteit Amsterdam, De Boelelaan 1118, Amsterdam, The Netherlands; Department of Neurology and Alzheimer Center Amsterdam, Amsterdam UMC location VUmc, De Boelelaan 1117, Amsterdam, The Netherlands; Department of Pathology, Amsterdam UMC location VUmc, De Boelelaan 1117, Amsterdam, The Netherlands; Netherlands Institute for Neuroscience, Meibergdreef 47, Amsterdam, The Netherlands; Amsterdam Neuroscience, Neurodegeneration, Amsterdam, The Netherlands; Delft Bioinformatics Lab, Delft Technical University, Van Mourik Broekmanweg 6, Delft, The Netherlands; Department of Clinical, Neuro and Developmental Psychology, Faculty of Behavioural and Movement Sciences, Vrije Universiteit Amsterdam,De Boelelaan 1118, Amsterdam, the Netherlands

**Keywords:** Centenarians, amyloid-beta, aging, Alzheimer’s disease, neuropathology, quantitative pathology, QuPath, *APOE*, cognition

## Abstract

**BACKGROUND:** Amyloid-beta(Aβ)-plaques accumulate in non-demented individuals, particularly at advanced ages. The unclear association between Aβ-pathology and cognition in elderly raises the question whether Aβ-pathology should be considered a benign consequence of aging.

**METHODS:** Post-mortem brains of 95 centenarians and 27 Alzheimer’s disease(AD) patients were evaluated for Aβ-plaque distribution according to the Thal phase and quantitative Aβ-load in the neocortex. For centenarians, Aβ-pathology was correlated to *APOE*-genotype and performance on 12 cognitive tests administered shortly before death.

**FINDINGS:** While 35% of centenarians exhibited Aβ-loads similar to AD patients, cortical Aβ-load was limited in 65% of centenarians, some of which had the highest Thal phase. Cortical Aβ-load, as opposed to Thal phase, associated with *APOE*-genotype and cognitive performance in centenarians.

**DISCUSSION:** Despite increasing Aβ-accumulation in various brain regions with age, actual Aβ-loads remain low in cognitively healthy centenarians. Therefore, Aβ-pathology in the oldest-old may not be considered a benign consequence of aging.

## 1. Back ground

Alzheimer’s disease (AD) is the most common cause of dementia, estimated to account for 60-80% of the 55 million dementia cases worldwide[1]. Clinically, AD is characterized by cognitive decline, including memory and learning impairment, and changes in behavior and personality. Neuropathological hallmarks of AD consist of: 1) amyloid-beta (Aβ) plaques; 2) hyperphosphorylated tau proteins accumulated as neurofibrillary tangles (NFTs) and; 3) neuritic plaques composed of fibrillary Aβ surrounded by dystrophic neurites[2]. The amyloid-cascade hypothesis postulates Aβ pathology as an initiator of downstream tau pathology, synaptic- and neuronal-loss, and eventually dementia[3]. Aβ plaques present in different forms, of which diffuse-, compact-, and cored-plaques are most common in sporadic AD[4]. Besides plaques, Aβ pathology can also accumulate in the vessel walls as cerebral amyloid angiopathy (CAA), and as subpial amyloid deposition[4, 5]. The widely accepted standard for assessing the severity of Aβ pathology in the post-mortem brain is the Thal Aβ phase system, which evaluates the extent of spatiotemporal Aβ distribution according to 5 phases, based on progressive deposition of Aβ in the neocortex (phase 1), allocortex (phase 2), diencephalon and striatum (phase 3), brainstem (phase 4), and the cerebellum (phase 5)[6]. Although there is a proposed order of involvement of different plaque-types at these different Thal phases, all Aβ plaque-types, but not CAA, are considered when assigning the Thal phase[4, 6].

In principle, AD patients are characterized by a Thal phase ≥3, while non-demented individuals aged <70 years generally exhibit little to no Aβ pathology (Thal phase ≤2)[6, 7]. However, with increasing age, Thal phase in non-demented cases gradually approaches the levels observed in AD patients[7]. As a matter of fact, we observed Thal phase 5 in the brains from centenarians, who were still cognitive healthy shortly before death[7, 8]. Accordingly, Thal phase does not associate with cognitive performance in cohorts with different ages, including centenarians[8, 9]. These observations previously led us to hypothesize that the deposition of Aβ plaques in self-reported cognitively healthy centenarians may be, at least in part, a natural consequence of aging and not directly causative of cognitive decline[8].

However, while the Thal phase reflects the hierarchical distribution of Aβ across brain regions, the actual load of Aβ pathology is not considered. As we observe great differences in Aβ-load in different brains with the same Thal phase, we questioned whether Aβ-load, rather than Thal phase, associates with cognitive performance close to death. Furthermore, we questioned whether Aβ-load in centenarians approaches the levels observed in AD patients with the same Thal phase. Moreover, we explored whether there is an effect of *APOE* genotype, the major genetic risk factor for AD, on Aβ-load in centenarians, as was previously described for younger populations[10–12].

To answer these questions, we analyzed postmortem brains from 95 centenarians and 27 AD patients. We evaluated Aβ pathology using; 1) the conventional Thal Aβ phase for the spatiotemporal distribution, and; 2) a quantitative imaging analysis method to determine Aβ-load in four neocortical regions, which are the first regions affected by Aβ pathology (i.e. in Thal phase 1). We compared Aβ pathology in centenarians to Aβ pathology in AD patients. Moreover, we explored the relation between the two different measurements for Aβ pathology. We furthermore investigated whether *APOE-*genotype associates with Aβ pathology in centenarians. Finally, we evaluated associations of Aβ pathology with cognitive functioning in centenarians, as measured close to death.

## 2. Methods

### 2.1 The 100-plus Study

The 100-plus Study enrolls self-reported cognitively healthy Dutch centenarians[13]. At baseline house-visit we: (1) collect lifetime history including educational history; (2) assess cognitive performance using a comprehensive test battery; (3) collect blood samples, allowing DNA isolation for genetic analysis including *APOE*-genotyping; and (4) discuss optional post-mortem brain donation. Cognitive performance in annually tested during follow-up visits. As a result of this, we have information on cognitive performance close to death. The 100-plus Study was approved by the local medical ethics committee of the VUmc (registration 2016.440) and all participants gave informed consent.

### 2.2 Cognitive assessment of centenarians

Cognitive assessment consisted of 16 tests addressing global cognitive performance and five cognitive domains (i.e. memory, verbal fluency, executive functions, visuospatial functioning and attention/processing speed). Cognitive test details are available in the Supplementary methods, and a detailed description of the cognitive testing procedures was published previously[13]. To minimize time between neuropsychological assessment and neuropathological evaluation, we included cognitive test scores collected at last study-visit. Some centenarians had missing test scores due to fatigue, sensory or motor difficulties, or because some tests were not yet included in the battery at the time of testing. Hence, we imputed missing test scores as described in the Supplementary methods. For the association between Aβ pathology and cognition, we included the 12 tests for which performance was available for >50% of centenarians prior to imputation (Table 1; Supplementary Table S1). Moreover, we calculated a composite global cognition score by combining normalized z-scores on 11 individual tests (excluding the Mini–Mental State Examination (MMSE), since this is a separate measure for global cognition). We only included the 88 centenarians for whom >50% of tests was available prior to imputation.

**Table 1.**
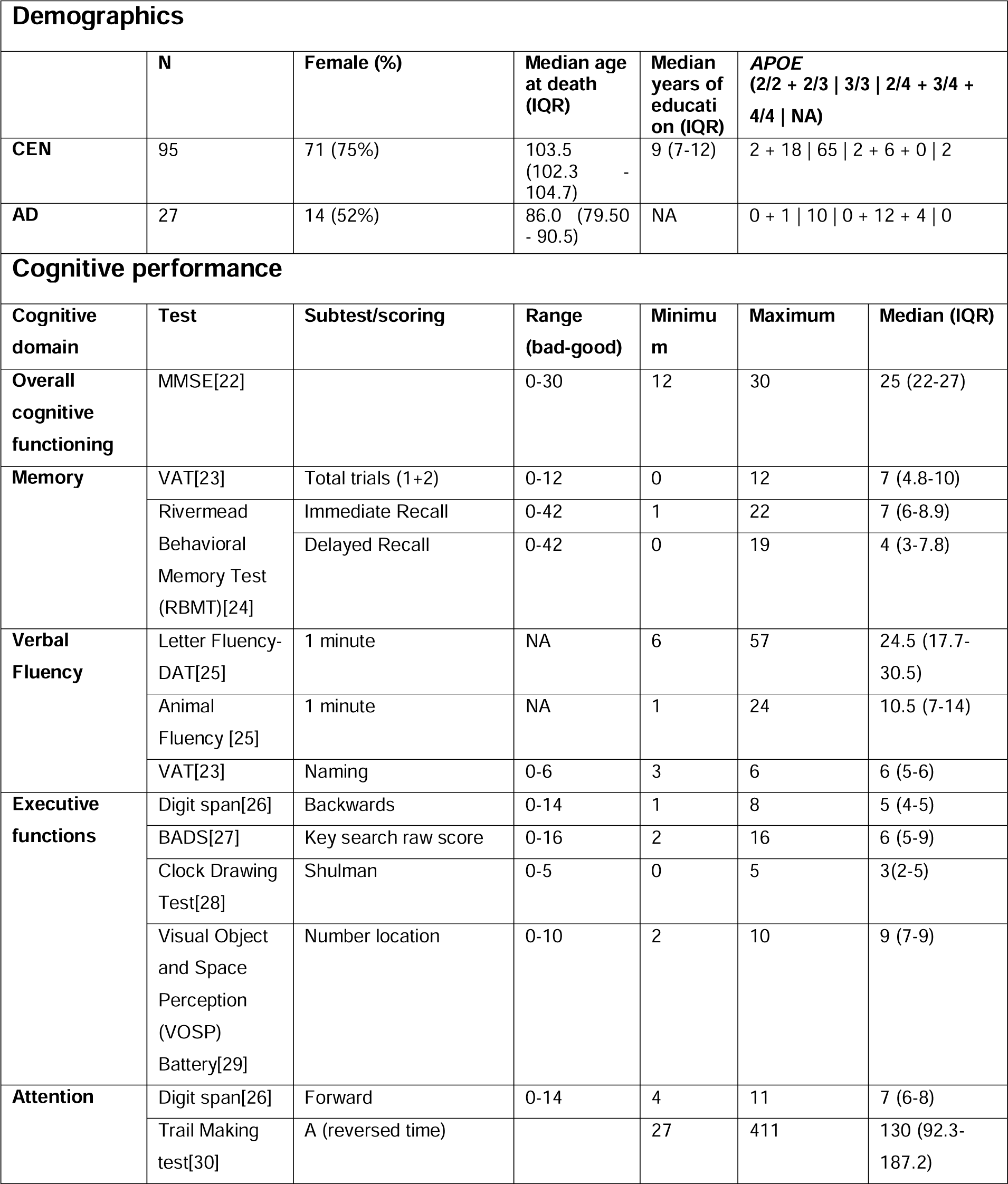
Demographics of the two cohorts and cognitive performance in the centenarian cohort. Overview of the cognitive domains and tests analyzed in relation to Aβ pathology in 88 centenarians. Minimum, maximum, interquartile range (IQR) and median test scores are given for the 88 centenarian cohort. Abbreviations: CEN= centenarians; AD= Alzheimer’s disease; IQR= interquartile range; NA = not available.

### 2.3 Post-mortem brain tissue

Post-mortem brain tissue from 95 centenarian-participants of the 100-plus Study and 27 AD patients were collected from 2012 until 2022. Brain autopsies were performed by the Netherlands Brain Bank (NBB; Amsterdam, The Netherlands (https://www.brainbank.nl), as approved by the local medical ethics committee of the VUmc (registration 2009.148). Donors granted informed consent for autopsy, tissue storage, and the utilization of anonymized clinical and neuropathological data for research. Brain donation was performed within 12 hours post-mortem, followed by formalin (10%) fixation for around 4 weeks. The dissection of brain tissue and neuropathological diagnosis followed the international guidelines established by the Brain Net Europe II consortium and NIA-AA, and were performed by one and the same neuropathologist[2, 14].

### 2.4 AD patients

As a reference for Aβ pathology in centenarians, we included late-onset AD patients from the NBB collection. We selected AD cases based on the criteria: 1) diagnosis of dementia during life; 2) intermediate or high score for AD pathology according to the NIA-AA criteria; 3) age at death ≥75; 4) *APOE* genotype available; and 5) Aβ stained cortical regions of interest available for Aβ-load quantification (see sections below).

### 2.5 Immunohistochemistry to visualize Aβ

For all autopsied brains, the NBB provided 8 µm thick formalin-fixed paraffin-embedded (FFPE) brain sections stained for Aβ (6F/D3, DAKO, Glostrup, Denmark, #M0872, 1:125) raised against amino acids 8-17 of the Aβ protein. The immunohistochemistry protocol is described in the Supplementary methods and was maintained as constant as possible throughout a period of 10 years.

### 2.6 Thal phases for spatiotemporal Aβ deposition

The spatiotemporal distribution of Aβ pathology was assessed using the Thal phase system, which ranges from 1 to 5[6]. Presence of Aβ plaques was evaluated in specific brain regions: middle frontal gyrus, middle temporal cortex, temporal pole cortex, inferior parietal lobule cortex, occipital pole cortex (if any number of Aβ plaques of any kind were observed in any of these regions, but none of the following, a Thal phase of 1 was assigned); CA1 region of the hippocampus and entorhinal cortex (Thal phase 2); molecular layer of the hippocampus, amygdala, nucleus caudatus, putamen and nucleus accumbens (Thal phase 3); CA4 region of the hippocampus, substantia nigra and inferior olivary nucleus (Thal phase 4); cerebellum and locus coeruleus (Thal phase 5). If no Aβ plaques of any kind were observed, Thal phase 0 was assigned. Neuropathological evaluation was done by one and the same neuropathologist.

### 2.7 Quantitative cortical Aβ-load

For each brain donor, we selected Aβ-stained sections of the middle frontal gyrus, inferior parietal lobe, temporal pole, and occipital pole (primary visual cortex), which are all involved in Thal phase 1 (Figure 1A). Slides were scanned at 20x magnification with an Olympus VS200 slide scanner and VS200 ASW software (version 3.3)(Figure 1B). Images (average size of 20 by 26 mm (± SD 2.3 and 2.9)) were analyzed in the open-source QuPath software (version 0.3.2)[15]. The grey matter was manually annotated as regions of interest (ROI)(Figure 1C). An artificial neural network was trained with a set of 10 images representing the full spectrum of Aβ immunoreactivity in terms of intensity and frequency. This pixel classifier was then applied to all images, where after any false negative and positive detections were used to refine the pixel classifier. Ultimately, a final pixel classifier accurately detected various Aβ immunoreactivity subtypes with varying intensities (Figure 1D-H). For each image, we calculated the Aβ-load as the percentage of the grey matter positive for Aβ immunoreactivity. Moreover, we binary classified images with an Aβ-load of 0% as negative.

**Figure 1.**
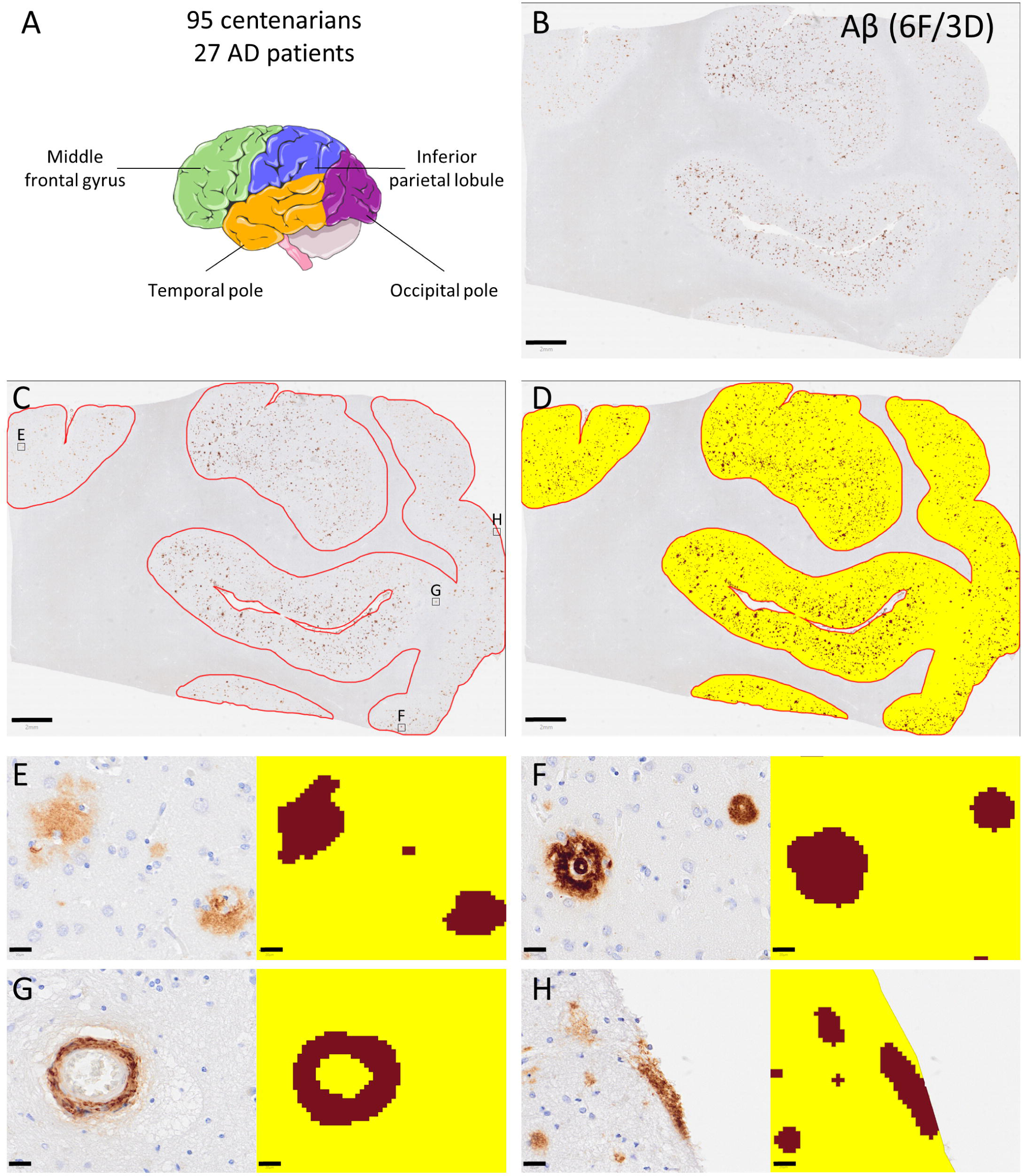
Quantitative cortical Aβ-load. For 95 centenarians and 27 Alzheimer’s disease (AD) patients, we analyzed the middle frontal gyrus, inferior parietal lobe, temporal pole and occipital pole (A). Digital images immunohistochemically stained for Aβ using the 6F/3D antibody in (B) were manually annotated for grey matter as the region of interest (C). Hereafter, a pre-trained pixel classifier was applied to detect Aβ immuno-positivity (D). The background and Aβ immuno-positivity are annotated with yellow and brown, respectively. Aβ-load was quantified as the percentage of cortex positive for Aβ (D). Aβ-load percentages included all Aβ positive deposits with a minimal diameter of 10 µm, mainly consisting of diffuse Aβ plaques (E), cored and compact-Aβ plaques (F), cerebral amyloid angiopathy (CAA) (G) and subpial amyloid deposition (H). Scale bars correspond to 2mm (B-D) and 20µm (E-H). This is an example image from the frontal cortex of a centenarian with an Aβ-load of 3.9% and a Thal phase of 5.

### 2.8 *APOE*-genotyping

Because of the low frequency of certain genotypes amongst centenarians, *APOE*-genotypes were grouped as: protective (ε2/2 and ε2/3), neutral (ε3/3), or risk-increasing (ε2/4 and ε3/4 (ε4/4 did not occur)[10].

### 2.9 Statistical analysis

Differences in median Thal phase and Aβ-load among cortical regions, cohorts or carriers of different *APOE-*genotypes, were tested with Kruskal-Wallis tests. The correlation between Thal phase and Aβ-load in various cortical regions was determined by calculating Spearman correlation coefficients. To examine different distributions of Aβ-negativity in specific brain regions or among different *APOE-* genotypes, we applied Fisher’s exact test and Chi-squared tests. To investigate the relation between Aβ pathology (predictors) and cognitive test scores (outcome), we applied multiple linear regression models. To ensure comparability of regression coefficients across different comparisons, all variables were normalized using z-scores. Models were corrected for the covariates sex, age at death, and years of education. All p-values were corrected for multiple testing by calculating the false discovery rate (FDR) using the Benjamini-Hochberg method. P-values <0.05 were considered significant. Analyses were performed in Rstudio (version 4.2.1.) and GraphPad Prism (version 9.3.1).

## 3. Results

### 3.1 Majority of centenarians had limited cortical Aβ-load

Thal phases in the centenarians were significantly lower (median 3, range 0-5) compared with AD patients (median 5, range 3-5) (p<1×10^−4^) (Figure 2A; demographics in Table 1). Across all regions, Aβ-load in centenarians was significantly lower compared to AD patients (p<1×10^−4^) (Figure 2B; see Figure S1 for corresponding images of Aβ immunopositivity in the frontal cortex). While it was not possible to quantify fractions of different types of Aβ depositions in the total Aβ-load, we observed that Aβ-load mainly represented diffuse-, cored-, and compact-Aβ plaques, and to a lesser extent CAA and subpial amyloid deposition (Figure 1E-H). We observed a spatiotemporal pattern of Aβ-load in the cortex in both centenarians and AD patients: Aβ-load was the highest in the frontal cortex, followed by the parietal cortex, then the temporal and occipital cortices (Figure 2B). In centenarians, this was also reflected in higher frequency of complete absence of Aβ pathology (i.e., 0% positive area) in the temporal and occipital cortices compared to parietal and frontal cortices (X^2^ (df=3)=7.8, p=0.04) (Table 2). Cortical Aβ-load correlated with Thal phase in centenarians, but not in AD patients as these were predominantly Thal phase 5 (Figure 2C). In all 33 centenarians with Thal phase 1-2, Aβ-load was <4% across all cortical regions (Figure 3A). In centenarians with Thal phases ≥3, we observed a wide variation in Aβ-load. In contrast, in AD patients we rarely observed cortical regions with Aβ-load <4%, and we never observed Aβ-load <4% across all cortical regions. Moreover, in the AD cohort, ∼4% was the lower limit of Aβ-load in the frontal cortex, the most affected region. Hence, we had multiple indications to divide the centenarian cohort into 62 (65%) centenarians with limited Aβ-loads (<4% Aβ-load across all regions) and 33 (35%) centenarians with AD-associated Aβ-loads (>4% Aβ-load in the frontal cortex) (Figure 4A). Intriguingly, 15 of the 30 centenarians with Thal phase 3 (50%), 3 of the 14 centenarians with Thal phase 4 (21%), and 2 of the 9 centenarians with Thal phase 5 (22%) were resistant to AD-associated levels of Aβ-load (Figure 4A-B). Overall, the median Aβ-load in frontal, parietal and occipital cortex was lower in centenarians compared to AD patients, even though they had Thal phase 4 and 5 (Figure 3B).

**Figure 2.**
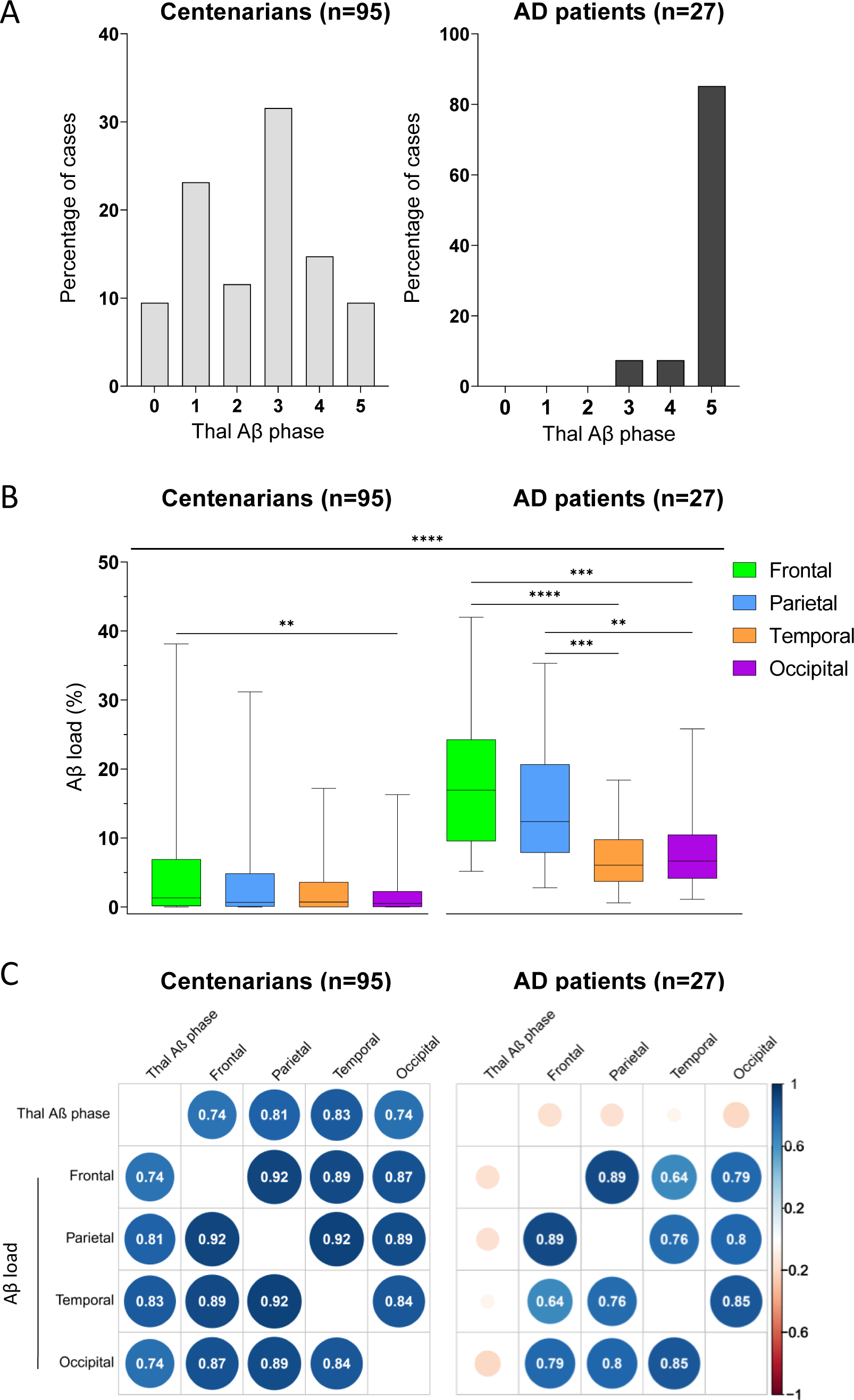
Centenarians have lower Thal phase and cortical Aβ-load compared to AD patients. (A) Density distribution of the Thal phases in the centenarian and AD cohorts. (B) Cortical Aβ-load (%) in the frontal, parietal, temporal and occipital cortices in centenarians versus AD patients. Kruskal-Wallis tests were used to compare Aβ-load in different regions within and between cohorts. Boxplots illustrate median, interquartile range and the minimum and maximum observed values. Asterisks indicate significance differences, with p-values (* ≤0.05, ** ≤0.01, and *** ≤0.001, **** ≤0.0001). See Figure S1 for images of Aβ immunopositivity in the frontal cortex representing the median and interquartile range in centenarians compared to AD patients. (C) Spearman correlations between Aβ pathology measurements. Color, size of the circles indicates the strength of the Spearman correlation coefficient, which is also given in each cell. Only significant correlations are given (p<0.05). For all graphs, p-values were corrected for false discovery rates (FDR) using Benjamini & Hochberg method.

**Figure 3.**
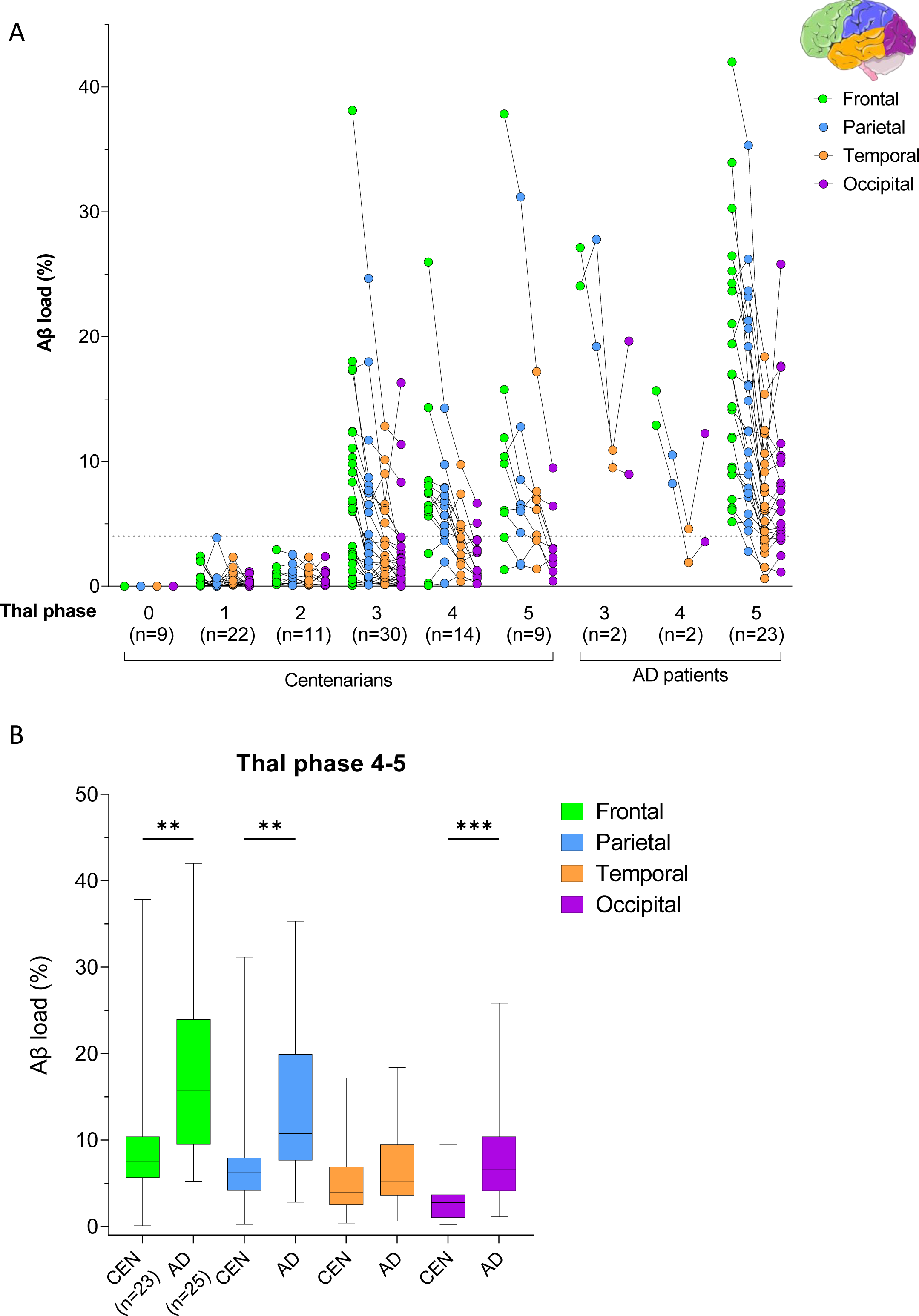
The relation between Thal phase and cortical Aβ-load. (A) Aβ-load (%) in the frontal, parietal, temporal and occipital cortices per Thal phase and per cohort. For each brain donor, the Aβ-load percentages in the different cortices are connected with a line. (B) Lower Aβ-load in frontal, parietal and occipital cortices from centenarians with Thal phase 4-5 compared to AD patients with Thal phase 4-5. Boxplots illustrate median, interquartile range and the minimum and maximum observed values. Kruskal-Wallis tests were used to compare Aβ-load between cohorts, and asterisks indicate the significance with p-values (* ≤0.05, ** ≤0.01, and *** ≤0.001). All p-values were corrected for false discovery rates (FDR) using Benjamini & Hochberg method.

**Figure 4.**
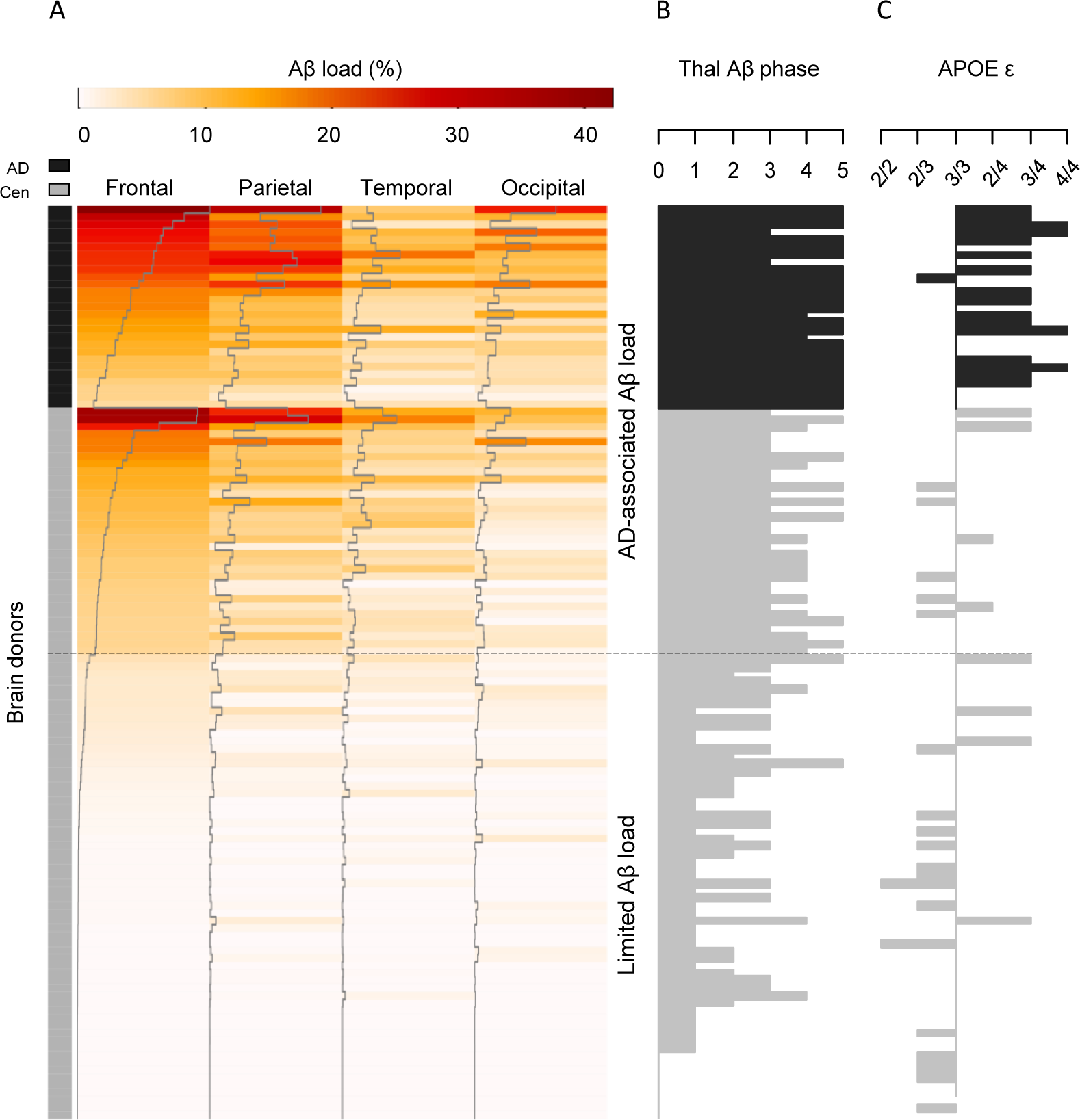
Cortical Aβ-load and corresponding Thal Aβ phase and *APOE* genotype in centenarians and AD patients. Each row represents one brain donor (centenarians n=95, AD n=27), sorted on Aβ-load in the frontal cortex. (A) The colors and the grey trace line of the heatmap represent cortical Aβ-load in frontal, parietal, temporal and occipital cortices. The Aβ-load heatmap demonstrates the strong inter-correlation between Aβ-load in different regions, with the frontal cortex most affected and the temporal and occipital cortices least affected. Based on the maximum observed Aβ-load in Thal phase 1-2 centenarians (<4%) and the Aβ-load observed in the frontal cortex of AD patients, we divided the cohort into a subset of 62/95 (65%) centenarians with limited Aβ-load and a subset of 33/95 (35%) with AD-associated levels of Aβ-load (dashed line). (B) Corresponding Thal phases are plotted. This demonstrated the overall correlation between Aβ-load and Thal phase, but also illustrates there are centenarians with Thal phase ≥ 3 but limited Aβ-load. (C) Corresponding *APOE* genotypes (missing for two cases) are plotted. A group of *APOE* ε2 carriers is enriched at the bottom of the heatmap, but five ε2 carriers (25%) had AD-associated levels of Aβ-load. Interestingly, half of the ε4-centenarians had limited Aβ-load, indicating they were resilient to the putative increasing effect of *APOE* ε4 on Aβ-load, as observed at the group level.

**Table 2.** Aβ negativity in the centenarian cohort. The number and percentage of centenarians presenting no immunoreactivity for Aβ (i.e. Aβ-load 0%) per brain region and in the entire brain (i.e. Thal 0) for the centenarian cohort as a whole, and per *APOE* group. Percentages were calculated over the total number of cases in the (sub)group on the left. NA = not available.

|  | **N** | **Frontal** | **Parietal** | **Temporal** | **Occipital** | **All regions/Thal 0** |
| --- | --- | --- | --- | --- | --- | --- |
| **Total** | 95 | 9 (9%) | 15 (16%) | 23 (24%) | 18 (19%) | 9 (9%) |
| ***APOE*** |  |  |  |  |  |  |
| **$\epsilon 2/2 + 2/3$** | 20 | 3 (15.0%) | 5 (25%) | 10 (50%) | 9 (45%) | 5 (25%) |
| **$\epsilon 3/3$** | 65 | 4 (6%) | 8 (12%) | 10 (15%) | 7 (11%) | 2 (2%) |
| **$\epsilon 2/4 + \epsilon 3/4$** | 8 | 0 (0%) | 0 (0%) | 1 (13%) | 0 (0%) | 0 (0%) |
| **NA** | 2 | 2 (100%) | 2 (100%) | 2 (100%) | 2 (100%) | 2 (100%) |

### 3.2 *APOE*-genotype associates with cortical Aβ-load in centenarians

Centenarians with a protective *APOE-*genotype (ε2/2 and ε2/3) had a lower Aβ-load across all cortical regions, compared to centenarians with a neutral (ε3/3)- or risk-increasing (ε2/4 and ε3/4)-*APOE-*genotype (Figure 5). Furthermore, *APOE-*ε2/2 and -ε2/3 carriers were more likely to be completely free of Aβ pathology in temporal and occipital cortices compared to other genotypes (Fisher’s exact test; p<0.001; Table 2). Five of the seven genotyped centenarians who were negative for Aβ (i.e. Thal 0) were ε2-carriers. On the other hand, *APOE*-ε4 carriers tended to have a higher Aβ-load compared to ε3/3-carriers, although this difference did not reach statistical significance, likely due to the limited number of ε4-centenarians (p_frontal_=0.112; p_parietal_=0.059; p_temporal_=0.377; p_occipital_ =0.127). Nevertheless, half of the ε4-carriers had limited Aβ-load, indicating they were resilient to the assumed effect of *APOE*-ε4 on Aβ-load, as observed on group level (Figure 4C).

**Figure 5.**
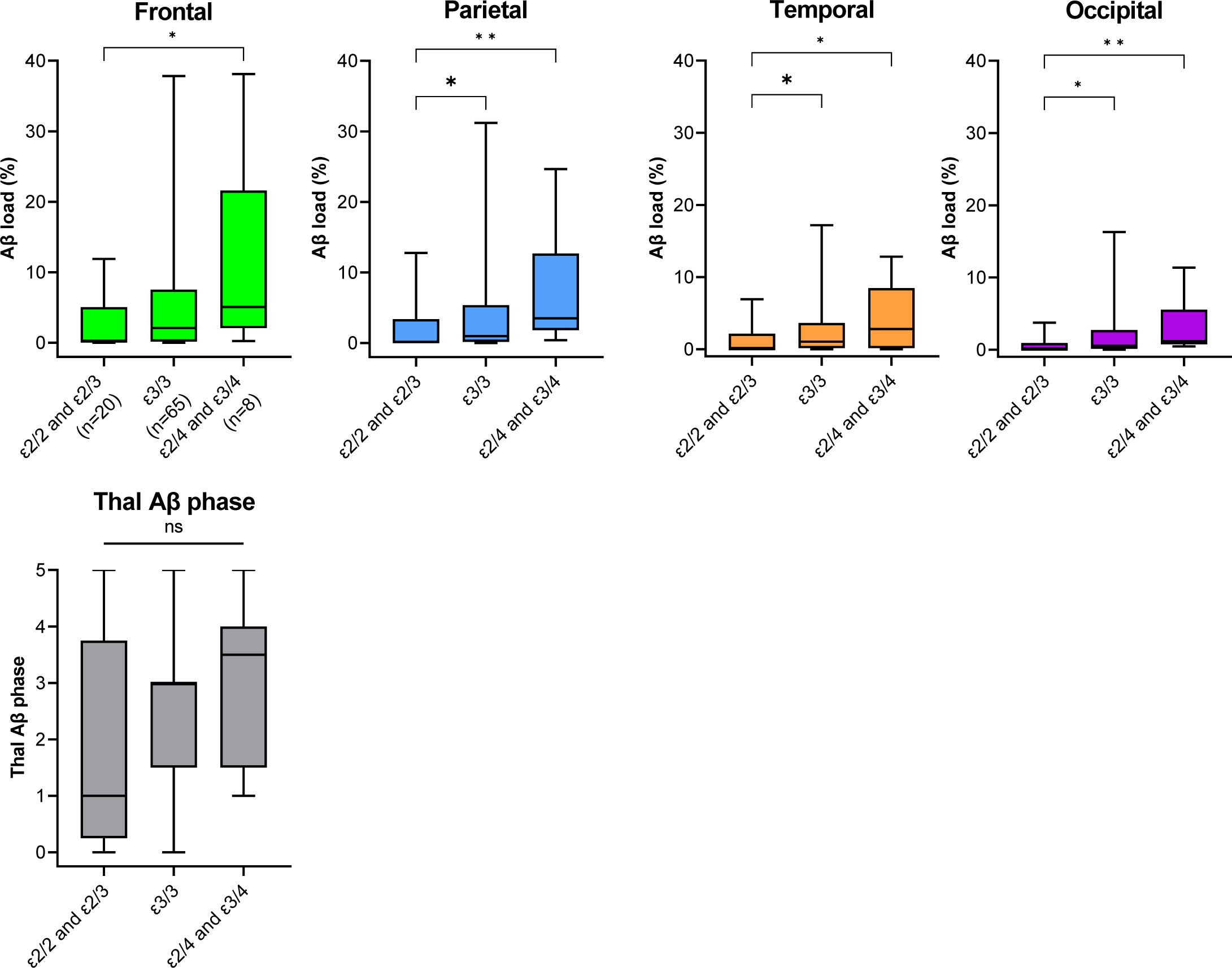
*APOE* genotype is associated with cortical Aβ-load in centenarians. Kruskal-Wallis tests were used to compare cortical Aβ-load and Thal phases between different *APOE* groups, with p-values (* ≤0.05, ** ≤0.01, and *** ≤0.001), corrected for false discovery rates (FDR) using Benjamini & Hochberg method.

### 3.3 Cortical Aβ-load affects global cognition and executive functioning in centenarians

For 88 centenarians, cognitive performance was assessed a median of 9.5 months prior to brain donation (interquartile range (IQR) 4-13) (Table 1). Regression models were applied to investigate the relation between Aβ pathology and cognitive test scores (Figure 6A, Table S2). Thal phase did not associate with cognitive performance. In contrast, higher Aβ-load associated with poorer global cognition, as measured with the MMSE and a composite score combining performance on all individual tests. Notably, higher Aβ-load across all regions associated with lower performance on the neuropsychological tests that assess executive functioning: the Digit Span backwards test, the Key search test, and the Clock Drawing test. Aβ-load did not associate with performance in other cognitive domains, indicating that the association with global cognition may be partly attributed to the executive functioning domain. In addition to the linear association, median performance on the aforementioned tests, except for the MMSE (p=0.075), was lower in the centenarian-group with AD-associated Aβ-load compared to the group with limited Aβ-load (p=0.075) (Figure 6B).

**Figure 6.**
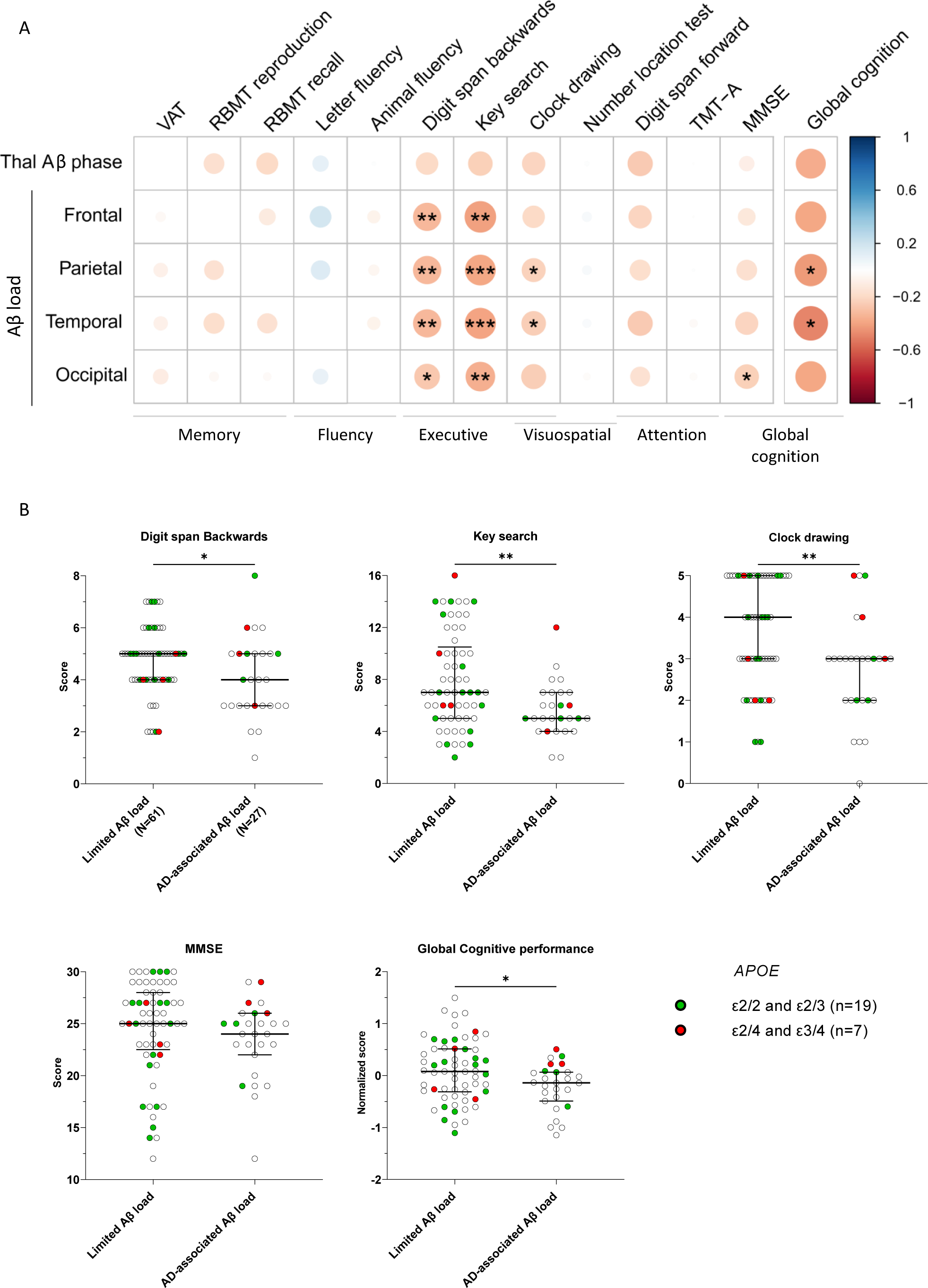
Cortical Aβ-load correlates with executive functioning and global cognition in centenarians. (A) Regression analysis between Aβ pathology (rows) and cognitive test scores (columns). Color and size of the circles indicate the strength of the regression coefficient, where blue indicates a positive correlation and red indicates a negative correlation. Analyses were corrected for covariates of age at death, sex (0 female, 1 male) and years of education (Table 1). p (* ≤0.05 and ** ≤0.01, ***≤0.001) corrected for false discovery rates (FDR) using Benjamini & Hochberg method. Regression coefficients and p-values can be found in Supplementary Table S2. (B) Median test score with interquartile range in the limited Aβ-load (n=61) group compared to the group with AD-associated levels of Aβ-load (n=27) (see Figure 4). Protective *APOE* genotypes are depicted in green (n=19), risk-increasing in red (n=7). Mann–Whitney U tests were used to compare test scores between the different groups. N=88. See Table 1 for details on cognitive performance.

Despite the strong associations between Aβ-loads and cognitive performance, individual signals were variable: some centenarians had AD-associated levels of Aβ-loads and scored high on neuropsychological tests, while others had a limited Aβ-load, but scored low on the cognitive tests (Figure 6B). This indicates that next to Aβ-load, additional factors associated with resilience may have affected individual cognitive performance. Although we observed a gene-dosage effect of *APOE-* ε2<ε3<ε4 on cortical Aβ-load (Figure 5), there was no association between *APOE-* genotype and cognitive performance in this group (Supplementary Figure S2). In fact, three ε4-centenarians with AD-associated Aβ-load maintained remarkably high levels of cognitive performance (Figure 6B).

## 4. Discussion

An analysis of Aβ pathology in 95 centenarians and 27 AD patients revealed that majority of centenarians resisted build-up of high levels of cortical Aβ-load typically observed in AD patients, even when Aβ spread was consistent with the highest Thal phases. Higher levels of cortical Aβ-load, but not higher Thal phase, associated with lower global cognitive performance and executive functioning. Furthermore, *APOE*-genotype associated with Aβ-load but not with Thal phase, further underscoring a role for Aβ-load, rather than Aβ distribution, in the risk of cognitive decline and AD. Overall, our findings suggest that keeping Aβ-load within limits is a hallmark of maintained cognitive health until extreme ages.

The finding that Aβ-load associates with cognitive performance in centenarians aligns with two previous reports that have linked post-mortem neocortical Aβ-load with lower cognition in non-demented 80-year-olds[16, 17]. Our study indicates that specifically the executive functioning domain, as assessed with the Digit Span Backwards, the Key Search, and the Clock Drawing tests, is affected by higher levels of Aβ-loads. Previous studies did not report a specific correlation with executive functioning, although one study reported a correlation between Aβ-load and the Digit Span Backwards test[17]. In line with our findings, a recent report indicated that increased Aβ-accumulation in the brain, as determined by cerebrospinal fluid or positron emission tomography (PET) measurements, specifically associated with lower executive functioning in non-demented individuals aged 50-80[18].

Note that in a previous analysis of this same set of centenarians we found that overall neuropathological changes, including NFT, TAR DNA-binding protein 43 (TDP-43), Lewy Bodies, and hippocampal sclerosis, associated strongest with the Clock Drawing test[8]. However, here we observe a stronger correlation between Aβ-load with the Key Search, and the Digit Span Backwards tests and to a lesser extent with the Clock Drawing tests. This may reflect sensitivity of the CDT for other neuropathological substrates, next to Aβ-load, while the Key Search and Digit Span may be more sensitive to high Aβ-loads.

This study is the first to directly compare conventional Thal phase to Aβ-load quantifications in relation to cognition. We show that quantitative Aβ-load correlates with cognition, while Thal phase does not. Our results emphasize the need for a wide and standardized application of quantitative pathology in addition to traditional staging systems, as it gives a more objective and representative measurement of pathology severity[19].

In both the centenarian and AD cohorts, we observed a spatiotemporal pattern of cortical Aβ load, wherein the middle frontal gyrus is affected earliest and most severely and the temporal and occipital poles are affected later and to a lesser degree. Our observations differ from the initial Thal phase study, in which Aβ plaques in 6 brains with Thal phase 1 were most frequently seen in temporal and occipital cortex (5/6), followed by the frontal cortex (4/6), and least in the parietal cortex (2/6)[6]. On the other hand, our observations are in line with a large study on ∼3500 post-mortem brains, in which the middle frontal gyrus and inferior parietal lobe had higher Aβ plaque count compared to the superior temporal gyrus and the occipital cortex[20]. Notably, we observed a similar pattern in centenarians as in AD patients, indicating comparable etiopathogenesis of Aβ pathology in healthy aging and AD.

Centenarian-carriers of the protective *APOE-*ε2 genotype had lower levels of Aβ-load, while the few carriers of the *APOE-*ε4 risk allele tended to have a higher Aβ-load. Moreover, the *APOE-*ε2 genotype was enriched in the small subset of centenarians who were fully resistant to the accumulation of Aβ (i.e. Thal phase 0). Altogether, while the risk increasing effect of *APOE-*ε4 on AD has been reported to wane with increasing age, our results suggest that the gene-dosage effect of ε2<ε3<ε4 on cortical Aβ-load, as observed in younger individuals, extends until extreme ages[11, 12, 21].

Nevertheless, despite the overall trend, half of the ε4-centenarians resisted the accumulation of AD-associated Aβ-load levels. Further investigation of these ε4-carriers may uncover mechanisms underlying resistance to the well-known effect of the ε4-allele on increased Aβ-load[12]. The other ε4-centenarians had Aβ-load levels similar to AD patients, but maintained remarkably high cognitive performance. We anecdote one ε4-carrier, who presented with the highest Aβ-load in the centenarian cohort (Figure 4), but who, at last study visit 9.5 months prior to brain donation, scored 26/30 on the MMSE, 5/5 on the Clock Drawing test, and above cohort average on global cognition (z-score 0.23), while scores on the Key search test (4/16) and Digit span Backwards (3/8) were relatively low. This illustrates that it is possible to maintain relatively high cognitive abilities despite high levels of Aβ-load. Investigation of these centenarians may guide us towards mechanisms that protect against factors with a risk-increasing effect on AD.

One of the inclusion criteria for the longitudinal 100-plus Study is that centenarians self-report to be cognitively healthy, which is confirmed by a proxy. At 100 years, the dementia incidence is ∼40%[13], such that at last annual study visit, commonly 3-5 years after study inclusion, centenarians present with a range of cognitive abilities. Therefore, the 100-plus Study cohort provides a unique window of opportunity to compare the relationship between Aβ pathology in the post-mortem brain with cognitive performance assessed shortly before death. Given the inclusion criteria of the study, the cohort includes an overrepresentation of individuals who are resistant and/or resilient to risk factors of dementia, as compared to the average population. Furthermore, given their age, centenarians have accumulated diverse disease-associated neuropathological substrates which may have influenced cognitive performance, which we did not take into account. Moreover, we analyzed total Aβ-load without discriminating between different pathological structures such as classic cored plaques, diffuse plaques, CAA, or subpial depositions. While all plaque types and subpial depositions are considered for Aβ-load, CAA is excluded for Thal phasing and evaluated with a separate CAA-Thal phase[5]. While the contribution of CAA to the total Aβ load is expected to be limited, this should nevertheless be taken into account when comparing the Aβ-load with Thal phases. Lastly, future studies are needed to explore whether higher Aβ-load in brain regions other than the cortex may associate with decline in other cognitive domains than executive function. For example, it would be possible that hippocampal Aβ-load may associate with decline of memory function.

In conclusion, we show that resistance to cortical Aβ-load, rather than Aβ-distribution, is an important contributor to maintaining cognitive health up until extreme ages. We have shown that cortical Aβ-load is associated with cognitive performance, specifically in the executive domain, and is influenced by *APOE-*genotype. Nevertheless, some centenarians were able to maintain high levels of cognitive performance despite accumulating high levels of Aβ. Finally, our results indicate that Aβ-pathology is not a benign consequence of aging, which highlights the prospective impact of emerging therapies aimed at reducing Aβ levels.

## Data Availability

All data produced in the present study are available upon reasonable request to the authors

## Acknowledgements

Neuropathological evaluation was performed by A.J.M.R. and the NBB, supported by S.K.R. and J.J.M.H. Quantitative pathology analysis were performed by S.K.R. and P.F.H. Neuropsychological assessment and collection of data for the 100-plus Study were performed by M.G., M.v.d.H., D.D., and L.M.C.L. Data analysis was performed by S.K.R., supported by S.A.M.S., M.Z., L.M.C.L., and M.H. Underlying data was verified by S.K.R., P.F.H., J.J.M.H., and H.H. The research was supervised by H.H. and J.J.M.H. Design of the research involved S.K.R., J.J.M.H., and H.H. The 100-plus Study was initiated by H.H. and supported by P.S. Design of the neurocognitive testing procedures of the 100-plus Study was supported by S.A.M.S. All authors commented on previous versions of the manuscript. All authors read and approved the final manuscript. We thank and acknowledge all participating centenarians and their family members. Moreover, we would like to thank and acknowledge all other brain donors and the NBB staff for the great cooperation.

## Consent statement

The study protocol was approved by the Medical Ethics Committee of the Amsterdam UMC. Informed consent was obtained from all participants. Brain donors consented to brain donation. The study was conducted in accordance with the Declaration of Helsinki.

