## Supplementary material for "Resistance to cortical amyloid-beta associates with cognitive health in centenarians"

### Supplementary methods

#### Cognitive tests

Global cognitive performance was assessed with the Mini–Mental State Examination (MMSE)[1]. Memory was evaluated using the Consortium to Establish a Registry for Alzheimer's Disease (CERAD) 10-word list immediate and delayed recall, Visual Association Test (VAT) and Rivermead Behavioral Memory Test (RBMT) immediate and delayed recall[2-4]. Verbal fluency was measured using the D-A-T letter fluency and animal fluency tests[5]. Executive functioning was assessed with the Digit Span Backwards test, Key search task and Rule Shift Cards subtests from the Behavioural Assessment of the Dysexecutive Syndrome (BADS), Trail Making Test (TMT)-part B, and the Meander test from the Amsterdam Dementia Screening Test[6-9]. Visuospatial functioning was measured with the Clock Drawing Test, Number Location test from the Visual Object and Space Perception (VOSP) battery and figure copying from the Cambridge Cognitive Examination (CAMCOG)[10-12]. Attention/processing speed was assessed with the Digit Span Forward test and the TMT-A[6, 8]. For downstream analysis, we only included tests for which scores were available for >50% of centenarians (Supplementary Table S1). We calculated a composite global cognition score by combining normalized z-scores on all individual tests available for >50% of centenarians (Supplementary Table S1), except the MMSE).

### Missing cognitive test score imputation

For some brain donors, test scores at last visit were missing due to fatigue, sensory or motor difficulties, or because some tests were not yet included in the test battery at the time of testing. Hence, we imputed the missing cognitive test scores as described previously[13-15]. In short, we applied multiple imputation by chained equations (MICE; version 3.13.0), which was found to be effective for the imputation of missing cognitive test scores, using 10 iterations based on the predictive mean matching method[16, 17]. To improve imputation quality, the input data was extended to all 400 centenarians that were included in the study at the moment of data analysis, and included all collected last-visit test scores (i.e., including four tests for which last-visit scores were available from <50% of the centenarians (Supplementary Table S1)). Next to neuropsychological test scores, input variables included parameters that were previously shown to correlate with cognitive performance in centenarians: years of education, the last-visit score on the Barthel Index for Activities of Daily living (ADL), the baseline-visit score on the Dutch Adult Reading test (DART) for intelligence, and, if applicable, the age at death[15, 18, 19]. To verify quality of imputation, we compared pre- and post-imputation minimum, maximum and median test scores (Supplementary Table S1). For the association with Aβ pathology, we only included brain donors for which at least 50% of input values was available (n=88).

### Immunohistochemistry to visualize Aβ

Sections were deparaffinized and rehydrated in a series of xylene and ethanol. Antigen retrieval was performed by boiling in citrate buffer (pH 6.0) for 10 minutes, followed by rinsing with distilled water twice, and by incubating with 80% formic acid for 5 minutes. Thereafter, sections were rinsed in distilled water twice and in phosphate-buffered saline (PBS) once. The sections were incubated with the primary antibody diluted in

PBS containing bovine serum albumin (BSA) (1%) overnight at 4°C or for 1 hour at room temperature. Sections were rinsed thrice in PBS and incubated with EnVision (anti-mouse/rabbit HRP, DAKO, Glostrup, Denmark, #K5007) for 1 hour at room temperature and washed thrice. This was followed by treatment with 3,3'-diaminobenzidine (DAB) for +/- 10 minutes for visualization of Aβ in brown. Nuclei were counterstained with hematoxylin, followed by dehydration in a series of alcohol and xylene.

### Supplementary Figures

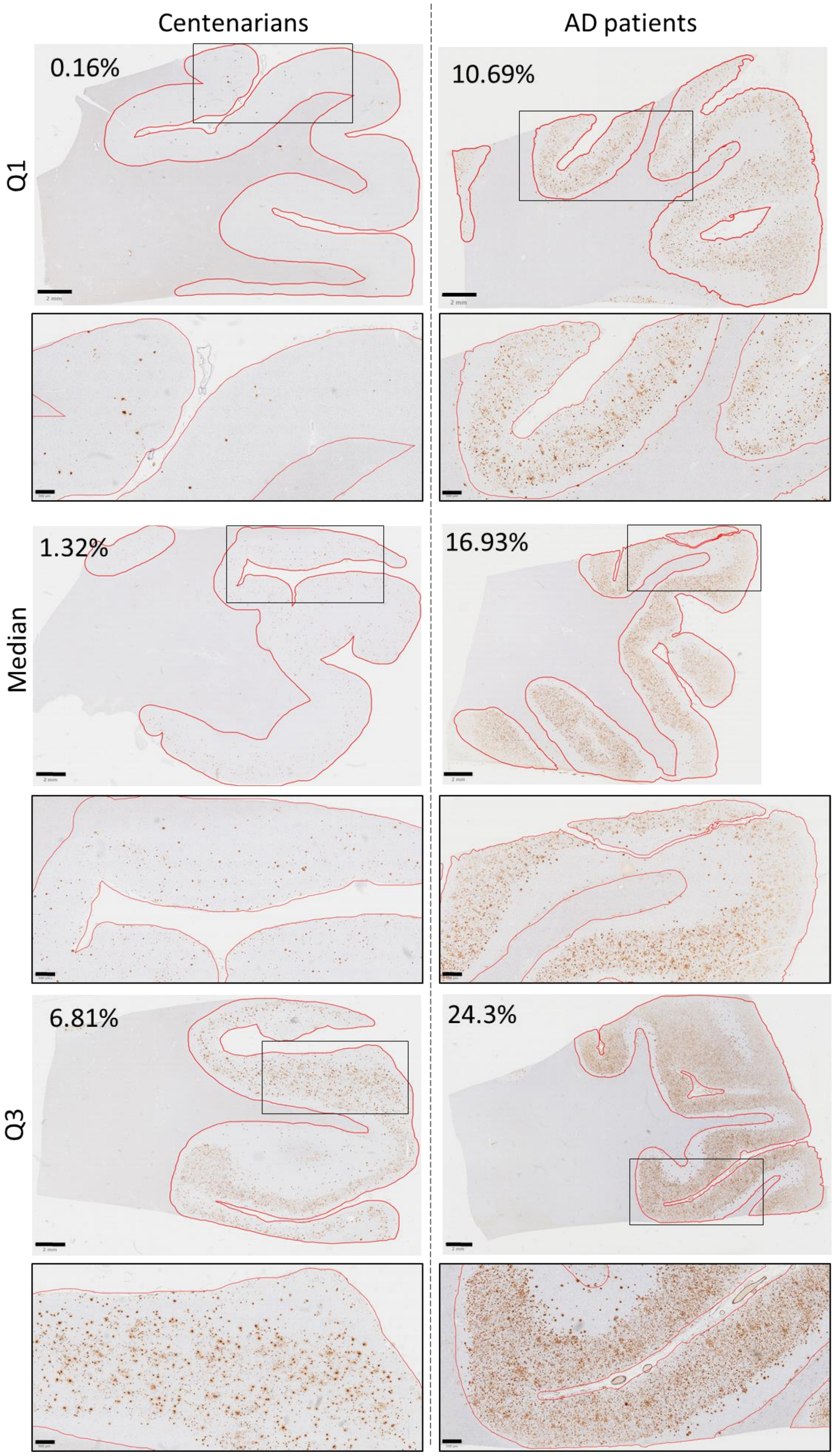
Centenarians
AD patients
Q1
0.16%
10.69%
Median
1.32%
16.93%
Q3
6.81%
24.3%

**Figure S1. Images representative of the first quartile, median and third quartile of Aβ-load in the frontal cortex for the centenarian and AD cohorts, illustrating lower Aβ-load in centenarians compared to Aβ patients.** Q1= first quartile, Q3= third quartile, AD= Alzheimer's disease. Scale bars correspond to 2mm and 500μm. Grey matter was annotated as the region of interest in red. Images correspond to Q1, median and Q3 values in the boxplot in Figure 2B.

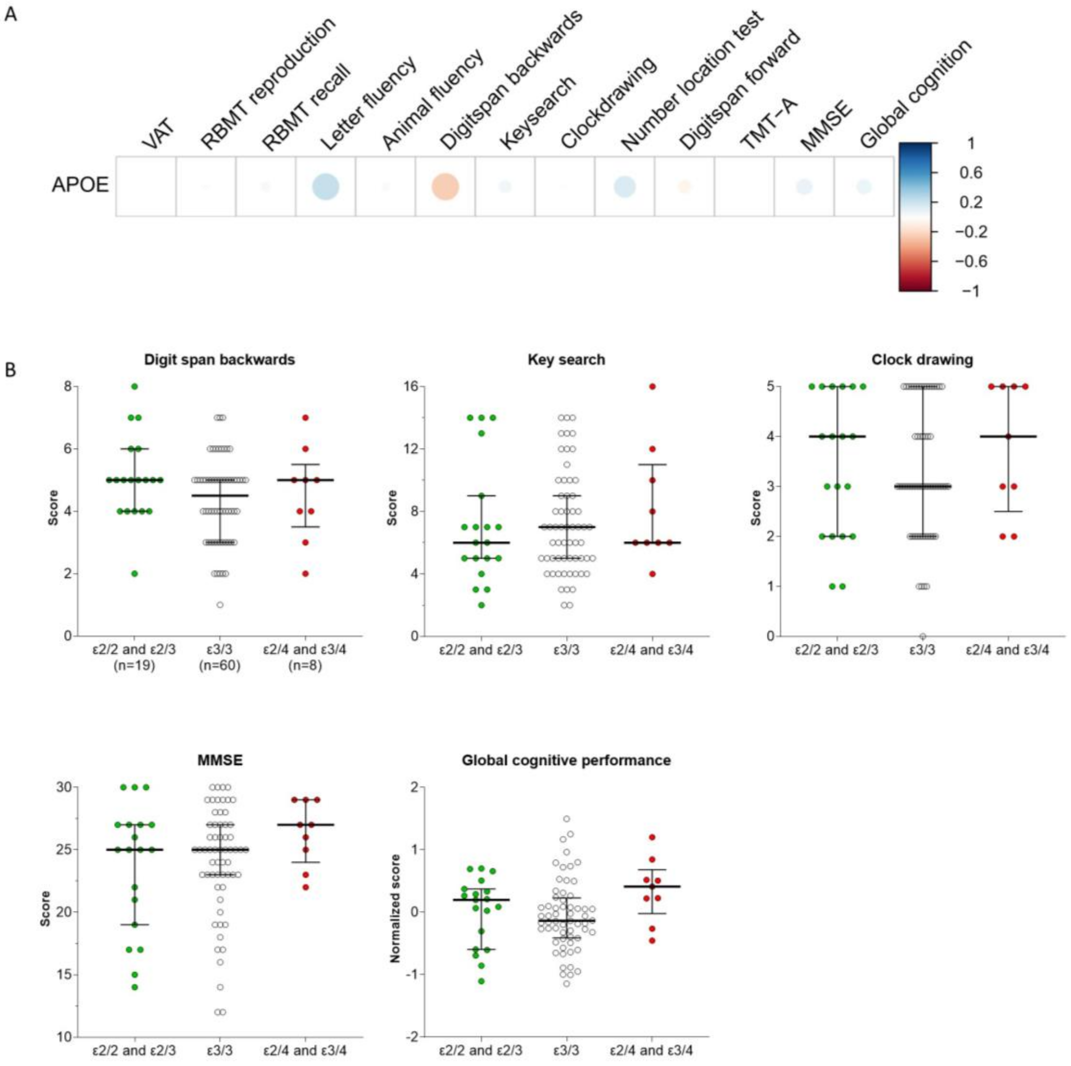

**Supplementary figure S2. No association between *APOE* genotype and cognitive performance in centenarians.** (A) Regression analysis between ordinal *APOE* genotype (protective = -1 (n=19), neutral =0 (n=60), risk increasing =1 (n=8)) and cognitive test scores. Color and size of the circles indicate the strength of the regression coefficient, where blue indicates a positive correlation and red indicates a negative correlation. Analyses were corrected for covariates of age at death, sex (0 female, 1 male) and years of education (Table 1). P-values were corrected for false discovery rates (FDR) using Benjamini & Hochberg method, there were no significant associations. (B) Median test score with interquartile range in the different *APOE*

groups. Kruskal-Wallis tests were used to compare test scores between the different groups. N=88. See supplementary Table 1 for details on cognitive performance.

**Supplementary Table S1. Input and output of the missing test score imputation.** Descriptive statistics of the input for cognitive score imputation, the output of the imputation (only tests for which >50% of scores were included) and the scores in the brain donor cohort (n=88) used for further analysis. IQR= interquartile range.

| Test details | | | | Pre-imputation (INPUT) | | | | Post-imputation (OUTPUT) | | | Brain cohort (n=88) | | |
|---|---|---|---|---|---|---|---|---|---|---|---|---|---|
| **Cognitive domain** | **Test** | **Subtest/scoring** | **Range (bad-good)** | **N (%)** | **Min** | **Max** | **Median (IQR)** | **Min** | **Max** | **Median (IQR)** | **Min** | **Max** | **Median (IQR)** |
| **Overall cognitive functioning** | MMSE[1] | | 0-30 | 400 (100%) | 8.4 | 30 | 24.8 (20.8-27.0) | 8.4 | 30 | 24.8 (20.8-27.) | 12 | 30 | 25 (22-27) |
| **Memory** | CERAD 10-word list[2] | Immediate Recall | 0-30 | 158 (40%) | 2 | 23 | 13.5 (11-17) | | | | | | |
| | | Delayed Recall | 0-10 | 153 (38%) | 0 | 10 | 3 (1-5) | | | | | | |
| | VAT[3] | Total trials (1+2) | 0-12 | 333 (83%) | 0 | 12 | 8 (4-11) | 0 | 12 | 7.6 (4-11) | 0 | 12 | 7 (4.8-10) |
| | Rivermead Behavioral Memory Test (RBMT)[4] | Immediate Recall | 0-42 | 237 (59%) | 0.5 | 28 | 7 (4.5-10.5) | 0.5 | 28 | 7.3 (5.5-9.5) | 1 | 22 | 7 (6-8.9) |
| | | Delayed Recall | 0-42 | 235 (58%) | 0 | 26 | 3.5 (1.5-7) | 0 | 26 | 3.8 (2.2-5.7) | 0 | 19 | 4 (3-7.8) |
| **Verbal Fluency** | Letter Fluency-DAT[5] | 1 minute | NA | 338 (85%) | 2 | 59 | 22 (14-29) | 2 | 59 | 22 (15-29) | 6 | 57 | 24.5 (17.7-30.5) |
| | Animal Fluency [5] | 1 minute | NA | 363 (91%) | 1 | 28 | 10 (7.5-14) | 1 | 28 | 10 (8-14) | 1 | 24 | 10.5 (7-14) |
| | VAT[3] | Naming | 0-6 | 304 (76%) | 0 | 6 | 6 (5-6) | 0 | 6 | 6(5-6) | 3 | 6 | 6 (5-6) |
| **Executive functions** | Digit span[6] | Backwards | 0-14 | 325 (81%) | 0 | 10 | 5 (4-5) | 0 | 10 | 4.3 (3.6-5) | 1 | 8 | 5 (4-5) |
| | BADS[7] | Key search raw score | 0-16 | 267 (67%) | 1 | 16 | 6 (4-9) | 1 | 16 | 6.2 (5-8) | 2 | 16 | 6 (5-9) |
| | BADS[7] | Rule shift Cards condition 2 (corrected profile score) | 0-4 | 120 (30%) | 0 | 4 | 1 (1-2.3) | | | | | | |
| | Trail Making test[8] | B (reversed time) | NA | 162 (41%) | 78 | 761 | 300 (206-357) | | | | | | |
| | Amsterdam Dementia Screening Test[9] | Meander | 0-4 | 102 (26%) | 0 | 4 | 4 (2-4) | | | | | | |

| | | | | | | | | | | | | | |
|---|---|---|---|---|---|---|---|---|---|---|---|---|---|
| **Visuospatial functioning** | CAMDEX-R/N CAMCOG)[12] | Figure copying, sum of 3 | 0-3 | 135 (34%) | 0 | 3 | 2 (1-2) | | | | | | |
| | Clock Drawing Test[10] | Shulman | 0-5 | 326 (81%) | 0 | 5 | 3 (2-5) | 0 | 5 | 3(2.2-5) | 0 | 5 | 3(2-5) |
| | Visual Object and Space Perception (VOSP) Battery[11] | Number location | 0-10 | 203 (51%) | 1 | 10 | 9 (8-10) | 1 | 10 | 8.7 (7.6-9) | 2 | 10 | 9 (7-9) |
| **Attention** | Digit span[6] | Forward | 0-14 | 335 (83%) | 3 | 12 | 7 (6-8) | 3 | 12 | 7 (6-8) | 4 | 11 | 7 (6-8) |
| | Trail Making test[8] | A (reversed time) | | 251 (62%) | 27 | 584 | 107 (77.5-160.5) | 27 | 584 | 125.5 (92.5-176) | 27 | 411 | 130 (92.3-187.2) |
| | Barthel Index[18] | | | 192 (48%) | 0 | 20 | 10.5 (6-15) | | | | | | |
| | Years of Education | | | 288 (72%) | 0 | 20 | 8 (6-11) | 0 | 20 | 9 (7-11) | 0 | 20 | 9 (7-12) |
| | Dutch Adult Reading test[19] | | | 285 (71%) | 13 | 100 | 74 (56.5-88) | | | | | | |
| | Age at death | | | 313 (78%) | 100.1 | 110.7 | 103.1 (101.9-104.5) | | | | 100.4 | 110.7 | 103.8 (102.3-104.7 |

**Supplementary Table S2. Regression coefficients and p-values for the relation between Aβ pathology and cognitive test scores.** Regressions were corrected for covariates of age at death, sex (0 female, 1 male) and years of education (Table 1). P-values were corrected for false discovery rates (FDR) using Benjamini & Hochberg method. Values correspond to Figure 6A.

| | **VAT** | | **RBMT reproduction** | | **RBMT recall** | | **Letter fluency** | | **Animal fluency** | | **Digit span backwards** | | **Key search** | | **Clock drawing** | | **Number location test** | | **Digit span forward** | | **TMT-A** | | **MMSE** | | **Global cognition** | |
|---|---|---|---|---|---|---|---|---|---|---|---|---|---|---|---|---|---|---|---|---|---|---|---|---|---|---|
| | **β** | **p** | **β** | **p** | **β** | **p** | **β** | **p** | **β** | **p** | **β** | **p** | **β** | **p** | **β** | **p** | **β** | **p** | **β** | **p** | **β** | **p** | **β** | **p** | **β** | **p** |
| **Thal Aβ phase** | -0.014 | 0.903 | -0.168 | 0.144 | -0.190 | 0.149 | 0.102 | 0.516 | 0.005 | 0.969 | -0.199 | 0.083 | -0.239 | 0.073 | -0.215 | 0.054 | 0.008 | 0.969 | -0.254 | 0.073 | -0.001 | 0.995 | -0.090 | 0.434 | -0.363 | 0.090 |
| **Frontal Aβ load** | -0.037 | 0.746 | -0.117 | 0.311 | -0.110 | 0.334 | 0.186 | 0.117 | -0.063 | 0.583 | -0.320 | **0.005** | -0.405 | **0.001** | -0.199 | 0.077 | 0.036 | 0.747 | -0.215 | 0.082 | -0.001 | 0.992 | -0.120 | 0.294 | -0.389 | 0.069 |
| **Parietal Aβ load** | -0.079 | 0.484 | -0.151 | 0.186 | -0.148 | 0.188 | 0.148 | 0.207 | -0.045 | 0.689 | -0.323 | **0.004** | -0.384 | **0.001** | -0.229 | **0.039** | 0.032 | 0.768 | -0.173 | 0.160 | 0.002 | 0.983 | -0.169 | 0.137 | -0.425 | **0.045** |
| **Temporal Aβ load** | -0.077 | 0.495 | -0.172 | 0.134 | -0.159 | 0.160 | 0.122 | 0.302 | -0.065 | 0.567 | -0.321 | **0.004** | -0.393 | **0.000** | -0.242 | **0.030** | 0.027 | 0.907 | -0.251 | 0.074 | -0.024 | 0.907 | -0.211 | 0.064 | -0.486 | **0.022** |
| **Occipital Aβ load** | -0.093 | 0.410 | -0.035 | 0.907 | -0.024 | 0.833 | 0.095 | 0.423 | -0.103 | 0.360 | -0.263 | **0.020** | -0.354 | **0.005** | -0.250 | 0.055 | -0.019 | 0.865 | -0.156 | 0.208 | -0.020 | 0.918 | -0.239 | **0.034** | -0.386 | 0.069 |
